# Accelerometer-Derived Physical Activity Volume, Not Pattern, Correlates with Reduced Gout Incidence in a UK Prospective Cohort

**DOI:** 10.1101/2025.08.24.25334337

**Authors:** Qiang Wang, Yueming Liu, Bin Zhu

**Affiliations:** Urology & nephrology Center, Department of nephrology, Zhejiang Provincial People’s Hospital, affiliated People’s Hospital, Hangzhou Medical College, Hangzhou, Zhejiang, China

## Abstract

**Background:** The role of physical activity (PA) patterns in gout prevention remains unclear. This prospective study investigated associations between accelerometer-derived PA patterns, specifically “regularly active” (RA) and “weekend warrior” (WW), and incident gout.

**Methods:** Among 97,474 gout-free UK Biobank participants with valid 7-day accelerometer data obtained using Axivity AX3 wrist-based triaxial devices, we categorized PA patterns as Inactive (<150 min/week moderate-to-vigorous physical activity [MVPA]), WW (≥150 min MVPA with ≥50% concentrated in 1–2 days) and RA (≥150 min MVPA, not meeting WW criteria). Multivariable Cox models were used to estimate hazard ratios (HRs) for incident gout.

**Results:** Over a median follow-up of 8.0 years, 905 incident gout cases occurred. Compared to the Inactive group (n=36,785), both RA (n=20,298; HR 0.75, 95% CI 0.62–0.91, P=0.004) and WW (n=40,391; HR 0.84, 95% CI 0.73–0.98, P=0.024) showed significantly reduced gout risk in fully adjusted models. Crucially, after additional adjustment for total MVPA volume, no significant difference in gout risk was observed between RA and WW across models (all P>0.3). Inverse probability of treatment weighting analyses confirmed these associations (RA vs. Inactive: HR 0.76, 95% CI 0.62–0.93, P=0.007; WW vs. Inactive: HR 0.85, 95% CI 0.72–0.99, P=0.039). Stratification by gout polygenic risk score tertiles showed consistent protective effects for both RA and WW patterns across all genetic risk strata (all P-interaction >0.1). Sensitivity analyses (varying MVPA thresholds, stricter WW definitions [≥75% MVPA in 1-2 days], excluding early gout cases) supported the robustness of the findings.

**Conclusion:** Physical activity, including both regularly active and weekend warrior patterns, was associated with significantly reduced gout risk. Physical activity volume, rather than its temporal pattern, drives the reduction in gout risk. When achieving equivalent MVPA volume, no significant difference exists between regularly active and weekend warrior patterns. The protective role of physical activity was consistent under different subgroups and genetic risk profiles, supporting its potential as a broadly applicable strategy for gout prevention.

## Introduction

Gout is a common metabolic arthritis characterized by the deposition of monosodium urate crystals within joints and surrounding tissues, triggering intense inflammation. In recent years, the global prevalence of gout has risen significantly, fueled by increasing obesity rates, dietary changes, and an aging population. It has become a major chronic disease posing a serious threat to public health [1]. Beyond causing severe joint pain and functional impairment that markedly reduces quality of life, gout is also strongly associated with an elevated risk of cardiovascular disease, chronic kidney disease, and all-cause mortality. Despite a well-understood pathophysiology and the availability of cost-effective treatments, gout management remains suboptimal for many patients [2].

Physical activity, a crucial modifiable lifestyle factor, has been extensively proven to confer protective effects against various chronic diseases, including cardiovascular disease [3], type 2 diabetes [4], and certain cancers [5]. Underlying mechanisms include improved insulin sensitivity, reduced systemic inflammation, and weight control [6]. However, whether physical activity can reduce the risk of gout, and whether different physical activity patterns—including evenly distributed versus concentrated activity— have discrepant effects on the incidence of gout, remains unclear due to a lack of high-quality prospective evidence. Consequently, while the 2020 American College of Rheumatology gout management guidelines recommend weight loss, they do not provide specific strategies for achieving it and notably omit explicit recommendations for physical activity [7].

Conventional epidemiological studies primarily assess physical activity via questionnaires. However, these subjective methods suffer from inherent limitations, including recall bias and reporting errors, and struggle to accurately capture key dimensions of daily activity such as intensity, frequency, duration, and temporal patterns [8–11]. Accelerometers represent the gold standard for objective physical activity measurement, enabling the continuous and precise recording of 24-h activity intensity, sedentary time, and detailed activity patterns [12]. The UK Biobank, a large-scale prospective population cohort study, has compiled extensive health information, biological samples, and genetic data. Critically, it also includes wrist-worn accelerometer data collected over 7 days in a large subset of participants, providing vast, objective physical activity data [13]. This resource offers an unprecedented opportunity to investigate the complex relationship between physical activity and gout risk comprehensively. Elucidating this relationship will not only help validate the role of physical activity in the primary prevention of gout but may also reveal which specific activity patterns (e.g., evenly distributed versus concentrated activity) are most effective for reducing gout risk. Such findings will provide high-level evidence to guide the development of targeted, personalized gout prevention strategies.

Therefore, leveraging the rich accelerometer data within the large prospective UK Biobank cohort, this study aims to prospectively investigate the independent association between objectively measured physical activity patterns and incident gout.

## Methods

### Study Population

This study utilized data from the UK Biobank, a large-scale prospective cohort that recruited over 500,000 participants aged 37 to 73 years between 2006 and 2010 [14]. Ethical approval was granted by the North West Multi-centre Research Ethics Committee (REC reference: 21/NW/0157) and the UK National Information Governance Board, with all participants providing written informed consent.

To assess the association between physical activity and the risk of incident gout, this study initially recruited 502,006 individuals from the UK Biobank. Those with valid wrist-worn accelerometer data were included (N=100,034). After excluding individuals with baseline gout diagnosis (N=2,560), 97,474 participants without prior gout and with complete accelerometer data were retained. Missing covariates were imputed using “mice” R package. The final analytical cohort comprised these 97,474 individuals (Figure S1).

### Accelerometer-derived physical activity

Physical activity was quantified using Axivity AX3 accelerometers worn on the dominant wrist for 7 consecutive days (100 Hz sampling; ±8g range) [13]. Data processing included gravity calibration, low-pass noise filtering, non-wear detection (≥60 consecutive inactive minutes) and 5-second epoch intensity classification, etc. Moderate-to-vigorous physical activity (MVPA) was identified via a validated machine-learning algorithm (accuracy=88%, κ=0.80) [15]. Daily MVPA (Field ID 40045) was used for analysis.

### Gout

Participants were considered to have gout if they met any of the following criteria: Self-reported diagnosis of gout (Field ID 20002 with code 1466); Hospital admission or death records (ICD-10 codes M10). Due to differences in follow-up frequency and intervals among participants, the study defined the first-recorded gout event during follow-up as the endpoint. Therefore, the observation was considered to end at the earliest of the following: occurrence of gout, death, or the last known follow-up date (October 31, 2022).

### Covariates

During enrollment in the UK Biobank study, participants’ characteristics including sex, age, ethnicity, smoking status, alcohol consumption, educational attainment, and income level were collected via touchscreen questionnaires. Additionally, trained staff measured participants’ height and weight during their assessment visit, with body mass index (BMI) subsequently calculated using the standard formula. The Townsend Deprivation Index (TDI), derived from participants’ residential postcodes, integrates area-level aggregate data on car ownership, unemployment, homeownership, and household overcrowding [16]. A healthy diet score (HDS, range: 0-5) was calculated based on self-reported weekly intake frequencies of vegetables, fruit, fish, and meat, with higher scores indicating healthier dietary patterns [17]. History of cardiovascular disease (CVD) (ICD-10 codes I42), chronic kidney disease (CKD) (ICD-10 codes N18), type 2 diabetes (T2DM) (ICD-10 codes E11), and hypertension (ICD-10 codes I10-I13, I15) was ascertained from corresponding fields.

### Statistical Analysis

Primary analysis: Multivariable Cox proportional hazards models estimated hazard ratios (HRs) for incident gout across physical activity patterns, using the inactive group as reference. Models were adjusted for age, sex, ethnicity, BMI, smoking, alcohol, HDS, TDI, household income, education level, and baseline history of CVD, CKD, T2DM, and hypertension. The regularly active group served as an additional comparator.

Sensitivity analyses: To evaluate robustness, we conducted five supplementary analyses:

1. Repeated primary analysis using the quartile-based thresholds (25th: 94.1 min; 50th: 211.1 min; 75th: 374.4 min) of MVPA duration. (2) Modified the WW criterion to require ≥75% of MVPA within 1–2 days. (3) Excluded participants who developed the gout within 2 years of wearing the monitoring device. (4) tested interactions between physical activity patterns and prespecified subgroup variables. Stratified analyses and multiplicative interaction terms (PA pattern × subgroup variable) evaluated effect modification by: sex, age group (<65 vs ≥65 years), BMI category, smoking status, education level, alcohol consumption, HDS strata, and baseline history of CVD, CKD, T2DM, and hypertension.

### Restricted cubic splines (RCS)

RCS with 5 knots (placed at default quantile positions: 0.05, 0.35, 0.65, and 0.95) were employed to model the dose-response relationship between MVPA and gout risk. Cox proportional hazards models were used to estimate HRs and 95% CIs. Likelihood ratio tests were performed to evaluate nonlinearity. The median MVPA level served as the reference point (HR = 1). Observations with MVPA exceeding the 95th percentile were further excluded to mitigate the influence of extreme values.

### Inverse Probability of Treatment Weighting (IPTW)

To evaluate the causal effect of different physical activity patterns on the risk of gout, we employed the IPTW method within the framework of the Average Treatment effect on the Treated (ATT). We compared each active group (WW and RA) separately with the inactive group. Propensity scores (PS) were estimated using a multivariable logistic regression model that included covariates potentially associated with both physical activity and the risk of gout. Under the ATT framework, individuals in the exposed (active) group were assigned a weight of 1, while individuals in the control (inactive) group were weighted by PS/(1−PS). Covariate balance before and after weighting was assessed using the standardized mean difference (SMD), with an SMD < 0.1 indicating adequate balance. Based on the weighted sample, we applied Cox proportional hazards models to estimate the HRs and 95% confidence intervals (CIs) for gout incidence across groups. In addition, we calculated the weighted incidence rates of gout (per 1,000 person-years) for each group to reflect the absolute risk.

### Genetic Susceptibility Analysis

The gout polygenic risk score (PRS) was obtained from the Polygenic Score Catalog (https://www.pgscatalog.org/) as PGS000199. This score comprises 29 single nucleotide polymorphisms (SNPs) identified through genome-wide association studies (GWAS) in individuals of European ancestry [18,19]. Full details of the selected SNPs are provided in Table S1. Using the Swiss Army Knife tool on the UK Biobank Research Analysis Platform, the individual PRS was calculated as follows: For each SNP, the number of effect alleles (0, 1, or 2) carried by each participant was multiplied by the SNP’s published effect size (β coefficient) derived from GWAS. The resulting weighted values for all 29 SNPs were then summed to yield the final PRS for each participant. Based on the calculated PRS distribution, participants were stratified into tertiles, resulting in three genetic risk groups: low genetic risk (lowest tertile), intermediate genetic risk (middle tertile), and high genetic risk (highest tertile).

### Software Implementation

All analyses were performed using R (Version 4.3.0). Primary packages included “survival” (for Cox models), “mice” (for multiple imputation), “cobalt” (for IPTW), “rms” (for restricted cubic splines), and “forestploter” (for forest plot). Statistical significance was defined as two-sided P<0.05.

## Results

### 1. Baseline Characteristics of the Study Population

A total of 97,474 participants were included in the final analysis. Based on World Health Organization’s physical activity guidelines [20], participants were categorized as Inactive (<150 min/week MVPA; n = 36,785), Weekend Warrior (WW; ≥150 min/week MVPA with ≥50% concentrated within 1–2 days; n = 40,391), and Regularly Active (RA; ≥150 min/week MVPA not meeting WW criteria; n = 20,298) (Table 1). Statistically significant differences (p < 0.001) were observed across multiple baseline characteristics. Compared to the Inactive group, both RA and WW participants were younger, had a lower proportion of females, lower obesity prevalence, healthier lifestyle and socioeconomic profiles, higher proportions of ideal Healthy Diet Scores, higher levels of college education, and lower prevalence of current smoking. Median total MVPA time differed significantly (p < 0.001), with the RA group reporting the highest (417.60 min/week), followed by the WW group (286.58 min/week), and the Inactive group the lowest (69.48 min/week).

**Table 1:** Baseline characteristics of the study population according to physical activity pattern. Participants were classified as Inactive (<150 min/week MVPA), WW (≥150 min/week MVPA with ≥50% concentrated within 1–2 days), or RA (≥150 min/week MVPA not meeting WW criteria). Values are presented as median (IQR) for continuous variables and n (%) for categorical variables. Differences between groups were assessed using Kruskal-Wallis test for continuous variables and Pearson’s Chi-squared test for categorical variables.

| Characteristic | Inactive<br>N = 36,785<sup>1</sup> | Regularly Active<br>N = 20,298<sup>1</sup> | Weekend Warrior<br>N = 40,391<sup>1</sup> | P-value<sup>2</sup> |
| --- | --- | --- | --- | --- |
| Age |  |  |  | <0.001 |
| Median (Q1, Q3) | 58 (50, 63) | 56 (49, 61) | 57 (50, 62) |  |
| Gender |  |  |  | <0.001 |
| Female | 24,444 (66.45%) | 10,534 (51.90%) | 20,942 (51.85%) |  |
| Male | 12,341 (33.55%) | 9,764 (48.10%) | 19,449 (48.15%) |  |
| Ethnicity |  |  |  | <0.001 |
| Non white | 1,421 (3.86%) | 683 (3.36%) | 1,087 (2.69%) |  |
| White | 35,364 (96.14%) | 19,615 (96.64%) | 39,304 (97.31%) |  |
| HDS category |  |  |  | <0.001 |
| Ideal | 14,777 (40.17%) | 8,809 (43.40%) | 16,973 (42.02%) |  |
| Medium | 17,800 (48.39%) | 9,402 (46.32%) | 19,278 (47.73%) |  |
| Poor | 4,208 (11.44%) | 2,087 (10.28%) | 4,140 (10.25%) |  |
| BMI |  |  |  | <0.001 |
| Underweight | 144 (0.39%) | 188 (0.93%) | 230 (0.57%) |  |
| Normal | 11,464 (31.16%) | 9,560 (47.10%) | 17,279 (42.78%) |  |
| Overweight | 14,941 (40.62%) | 7,938 (39.11%) | 17,130 (42.41%) |  |
| Obese | 10,236 (27.83%) | 2,612 (12.87%) | 5,752 (14.24%) |  |
| TDI |  |  |  | <0.001 |
| Median (Q1, Q3) | -2.45 (-3.80, -0.19) | -2.06 (-3.61, 0.54) | -2.57 (-3.90, -0.47) |  |
| Education |  |  |  | <0.001 |
| UnKnown | 3,996 (10.86%) | 1,357 (6.69%) | 2,907 (7.20%) |  |
| College | 13,132 (35.70%) | 10,080 (49.66%) | 18,983 (47%) |  |
| Other levels | 19,657 (53.44%) | 8,861 (43.65%) | 18,501 (45.80%) |  |
| Drinking status |  |  |  | <0.001 |
| Never | 1,447 (3.93%) | 543 (2.68%) | 904 (2.24%) |  |
| Previous | 1,192 (3.24%) | 565 (2.78%) | 910 (2.25%) |  |
| Current | 34,146 (92.83%) | 19,190 (94.54%) | 38,577 (95.51%) |  |
| Smoking status |  |  |  | <0.001 |
| Never | 20,343 (55.30%) | 11,802 (58.14%) | 23,854 (59.06%) |  |
| Previous | 13,213 (35.92%) | 7,258 (35.76%) | 14,176 (35.10%) |  |
| Current | 3,229 (8.78%) | 1,238 (6.10%) | 2,361 (5.85%) |  |
| Household income |  |  |  | <0.001 |
| Less than 18,000 | 6,862 (18.65%) | 2,868 (14.13%) | 5,319 (13.17%) |  |
| 18,000 to 30,999 | 9,669 (26.29%) | 4,597 (22.65%) | 9,462 (23.43%) |  |
| 31,000 to 51,999 | 10,327 (28.07%) | 5,743 (28.29%) | 11,584 (28.68%) |  |
| 52,000 to 100,000 | 7,896 (21.47%) | 5,331 (26.26%) | 10,744 (26.60%) |  |
| Greater than 100,000 | 2,031 (5.52%) | 1,759 (8.67%) | 3,282 (8.13%) |  |
| Total MVPA |  |  |  | <0.001 |
| Median (Q1, Q3) | 69.48 (28.01, 109.31) | 417.60 (296.45, 590.40) | 286.58 (209.99, 417.60) |  |
| CVD |  |  |  | <0.001 |
| No | 26,186 (71.19%) | 16,169 (79.66%) | 31,768 (78.65%) |  |
| Yes | 10,599 (28.81%) | 4,129 (20.34%) | 8,623 (21.35%) |  |
| CKD |  |  |  | <0.001 |
| No | 35,933 (97.68%) | 20,076 (98.91%) | 39,809 (98.56%) |  |
| Yes | 852 (2.32%) | 222 (1.09%) | 582 (1.44%) |  |
| T2DM |  |  |  | <0.001 |
| No | 34,920 (94.93%) | 19,858 (97.83%) | 39,468 (97.71%) |  |
| Yes | 1,865 (5.07%) | 440 (2.17%) | 923 (2.29%) |  |
| Hypertension |  |  |  | <0.001 |
| No | 25,190 (68.48%) | 15,744 (77.56%) | 30,782 (76.21%) |  |
| Yes | 11,595 (31.52%) | 4,554 (22.44%) | 9,609 (23.79%) |  |

<sup>1</sup>n (%)
<sup>2</sup>Kruskal-Wallis rank sum test; Pearson's Chi-squared test

Density plots (Fig. S2) revealed distinct weekly MVPA distribution patterns: RA participants maintained consistent activity throughout the week, whereas WW participants concentrated the majority of MVPA within 1–2 days, with substantially lower activity on remaining days.

### 2. Association Between Physical Activity Patterns and Gout Risk

Cox proportional hazards models evaluated the association between physical activity patterns and incident gout over a median follow-up of 8.0 years, during which 905 incident gout cases were documented. In Model 1 (adjusted for age and sex only), both RA (HR=0.55, 95% CI: 0.46–0.67; P<0.001) and WW (HR=0.64, 95% CI: 0.55–0.74; P<0.001) showed significantly reduced gout risk compared to the Inactive group (Fig. 1A). These associations persisted in Model 2 after additional adjustment for BMI, ethnicity, TDI, education level, alcohol consumption, smoking status, household income, and HDS (RA: HR=0.70, 95% CI: 0.57–0.85; P<0.001; WW: HR=0.78, 95% CI: 0.68–0.91; P=0.001). After further adjustment for baseline history of CVD, CKD, T2DM and hypertension (Model 3), the protective association remained significant for RA (HR=0.75, 95% CI: 0.62–0.91; P=0.004) and WW (HR=0.84, 95% CI: 0.73–0.98; P=0.024) (Fig. 1A). Notably, WW participants accumulated significantly lower total weekly MVPA than RA participants (mean ± SD: 340.7 ± 181.8 vs. 479.7 ± 263.6 minutes; P<0.001) (Fig. S3). Crucially, after additional adjustment for total MVPA duration, no significant difference in gout risk was observed between the RA and WW groups across all models (all P>0.3; Fig. 1B), indicating that the protective effect of the two physical activity patterns per se against gout risk did not differ significantly.

**Figure 1:**
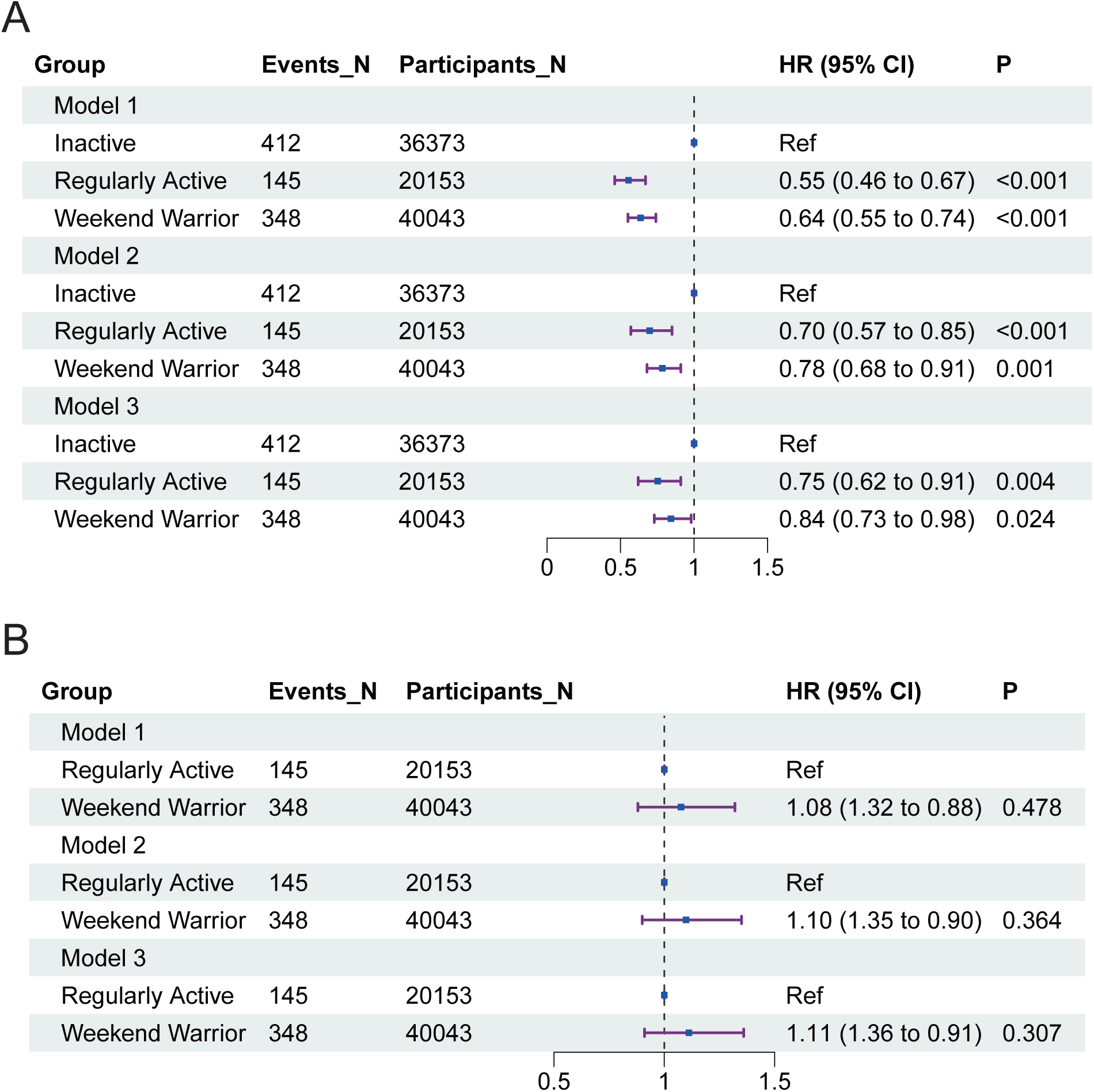
Association between physical activity patterns and gout risk. (A) Forest plot of HRs and 95% CIs for incident gout risk comparing WW and RA patterns to the Inactive group across Cox proportional hazards models (Model 1: age and sex adjusted; Model 2: additionally adjusted for BMI, ethnicity, TDI, education, alcohol, smoking, income, HDS; Model 3: further adjusted for baseline history of CVD, CKD, T2DM, hypertension). (B) Forest plot of HRs (95% CIs) comparing WW vs. RA patterns across the same models, after additional adjustment for total MVPA duration. P values for comparisons are indicated. The vertical dashed line represents HR = 1 (no effect). Ref, reference; HR, hazard ratio; CI, Confidence Interval; WW, Weekend Warrior; RA, Regularly Active; BMI, Body Mass Index; TDI, Townsend Deprivation Index; HDS, Healthy Diet Score; CVD, Cardiovascular Disease; CKD, Chronic Kidney Disease; T2DM, Type 2 Diabetes Mellitus; MVPA, Moderate-to-Vigorous Physical Activity.

Kaplan-Meier analyses corroborated these findings, showing significantly divergent cumulative gout incidence curves (overall log-rank P<0.001; Fig. 2). The Inactive group exhibited the highest cumulative incidence, while the RA group demonstrated the lowest. The WW curve occupied an intermediate position, overlapping substantially with RA. Pairwise comparisons (Bonferroni-corrected) confirmed significant risk reduction for both RA (vs. Inactive: P<0.001) and WW (vs. Inactive: P<0.001), but no statistically significant difference between RA and WW (P=0.157).

**Figure 2:**
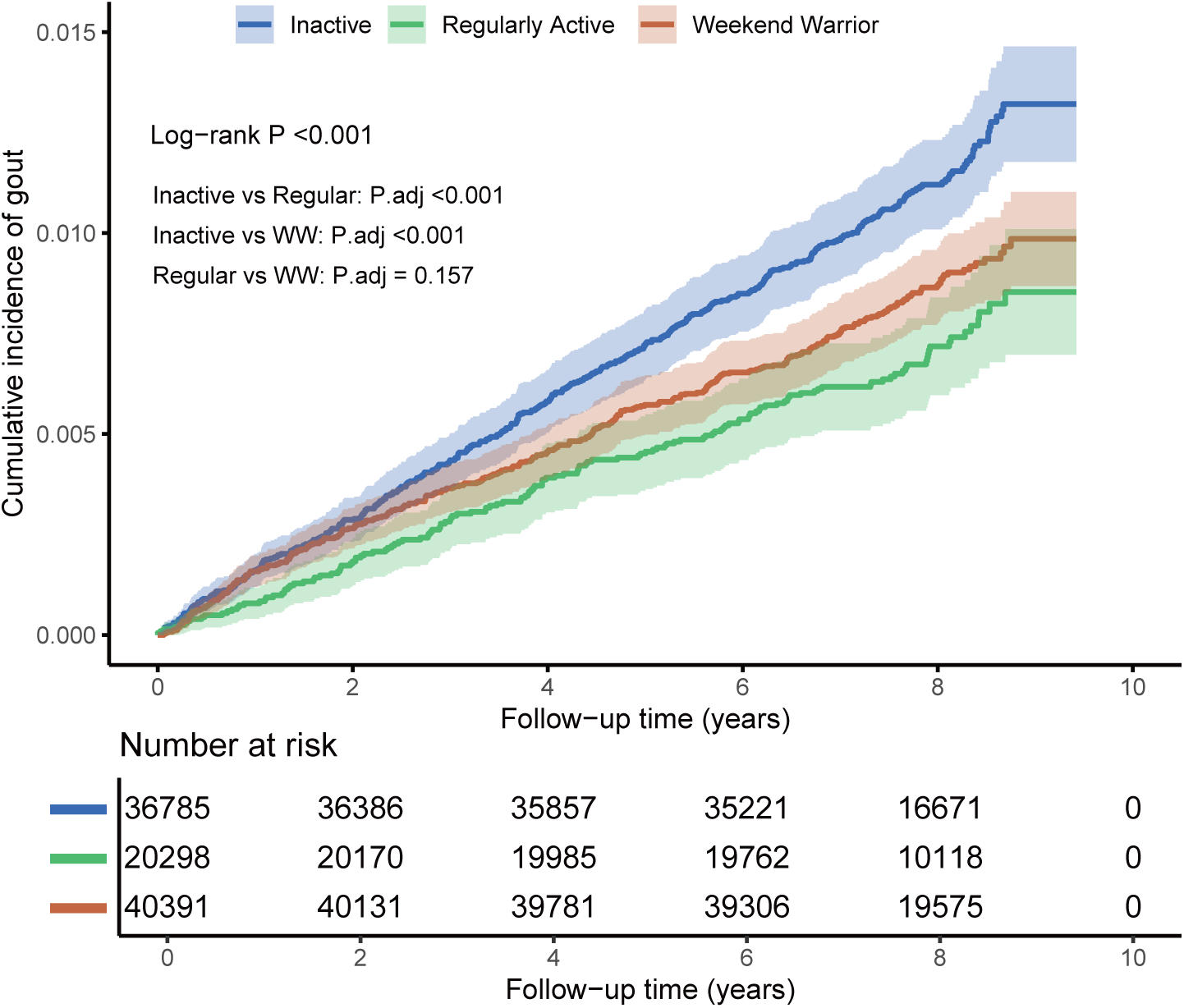
Cumulative incidence of gout according to physical activity pattern. Kaplan-Meier curves showing the cumulative incidence of gout over follow-up for Inactive, WW, and RA groups. Overall log-rank P < 0.001. Pairwise comparisons with Bonferroni correction confirmed significant differences between RA vs. Inactive (P < 0.001) and WW vs. Inactive (P < 0.001), but not between RA vs. WW (P = 0.157).

Subgroup analyses indicated that trends towards reduced gout risk were observed for both RA and WW patterns compared to inactivity (Table 2). Importantly, formal tests for interaction revealed no statistically significant effect modification by any subgroup variable (all P for interaction > 0.4), reinforcing that the inverse association between MPVA patterns and gout risk is robust across diverse population characteristics.

**Table 2:** Subgroup analyses of the association between physical activity patterns and incident gout risk. HRs and 95% CIs for gout risk comparing WW and RA patterns to the Inactive group. Estimates were derived from Cox proportional hazards models, adjusted for age, sex, BMI, ethnicity, TDI, education level, alcohol consumption, smoking status, household income, HDS, and baseline history of CVD, CKD, T2DM, and hypertension. Formal tests for interaction were performed for each subgroup variable.

| Subgroup | Variable | HR (95% CI) | P | P for interaction |
| --- | --- | --- | --- | --- |
| **Age** |  |  |  |  |
| ≥65 | Regularly Active | 0.75 (0.51-1.09) | 0.135 | 0.877 |
| <65 | Regularly Active | 0.73 (0.59-0.92) | 0.008 | 0.877 |
| ≥65 | Weekend Warrior | 0.88 (0.67-1.16) | 0.363 | 0.877 |
| <65 | Weekend Warrior | 0.83 (0.69-0.99) | 0.034 | 0.877 |
| **Gender** |  |  |  |  |
| Female | Regularly Active | 0.81 (0.52-1.25) | 0.338 | 0.418 |
| Male | Regularly Active | 0.74 (0.6-0.92) | 0.008 | 0.418 |
| Female | Weekend Warrior | 0.78 (0.56-1.08) | 0.139 | 0.418 |
| Male | Weekend Warrior | 0.86 (0.72-1.02) | 0.078 | 0.418 |
| **BMI** |  |  |  |  |
| Normal | Regularly Active | 1 (0.64-1.58) | 0.986 | 1.000 |
| Obese | Regularly Active | 0.63 (0.43-0.91) | 0.013 | 1.000 |
| Overweight | Regularly Active | 0.77 (0.58-1.01) | 0.062 | 1.000 |
| Underweight | Regularly Active | NA (NA-NA) |  | 1.000 |
| Normal | Weekend Warrior | 0.96 (0.65-1.44) | 0.858 | 1.000 |
| Obese | Weekend Warrior | 0.82 (0.64-1.05) | 0.109 | 1.000 |
| Overweight | Weekend Warrior | 0.85 (0.68-1.06) | 0.150 | 1.000 |
| Underweight | Weekend Warrior | NA (NA-NA) |  | 1.000 |
| **Education** |  |  |  |  |
| Other levels | Regularly Active | 0.71 (0.53-0.94) | 0.018 | 0.745 |
| College | Regularly Active | 0.79 (0.58-1.08) | 0.144 | 0.745 |
| Unknown | Regularly Active | 0.84 (0.5-1.41) | 0.498 | 0.745 |
| Other levels | Weekend Warrior | 0.8 (0.64-0.98) | 0.035 | 0.745 |
| College | Weekend Warrior | 0.85 (0.66-1.1) | 0.212 | 0.745 |
| Unknown | Weekend Warrior | 1.04 (0.71-1.53) | 0.827 | 0.745 |
| **Drinking status** |  |  |  |  |
| Current | Regularly Active | 0.77 (0.63-0.94) | 0.010 | 0.637 |
| Never | Regularly Active | 0.25 (0.03-1.96) | 0.187 | 0.637 |
| Previous | Regularly Active | 0.39 (0.08-1.84) | 0.235 | 0.637 |
| Current | Weekend Warrior | 0.84 (0.73-0.98) | 0.029 | 0.637 |
| Never | Weekend Warrior | 1.07 (0.42-2.73) | 0.890 | 0.637 |
| Previous | Weekend Warrior | 0.47 (0.14-1.57) | 0.221 | 0.637 |
| **Smoking status** |  |  |  |  |
| Never | Regularly Active | 0.81 (0.61-1.07) | 0.140 | 0.457 |
| Current | Regularly Active | 0.89 (0.44-1.8) | 0.746 | 0.457 |
| Previous | Regularly Active | 0.67 (0.5-0.9) | 0.008 | 0.457 |
| Never | Weekend Warrior | 0.84 (0.67-1.05) | 0.120 | 0.457 |
| Current | Weekend Warrior | 0.85 (0.48-1.5) | 0.565 | 0.457 |
| Previous | Weekend Warrior | 0.84 (0.68-1.05) | 0.120 | 0.457 |
| **HDS category** |  |  |  |  |
| Ideal | Regularly Active | 0.75 (0.53-1.05) | 0.093 | 0.993 |
| Poor | Regularly Active | 0.91 (0.58-1.44) | 0.693 | 0.993 |
| Medium | Regularly Active | 0.71 (0.54-0.94) | 0.017 | 0.993 |
| Ideal | Weekend Warrior | 0.84 (0.64-1.09) | 0.190 | 0.993 |
| Poor | Weekend Warrior | 0.8 (0.55-1.17) | 0.255 | 0.993 |
| Medium | Weekend Warrior | 0.86 (0.7-1.06) | 0.153 | 0.993 |
| **CVD** |  |  |  |  |
| Yes | Regularly Active | 0.74 (0.55-0.99) | 0.042 | 0.780 |
| No | Regularly Active | 0.76 (0.58-0.98) | 0.038 | 0.780 |
| Yes | Weekend Warrior | 0.91 (0.74-1.12) | 0.381 | 0.780 |
| No | Weekend Warrior | 0.79 (0.64-0.97) | 0.024 | 0.780 |
| **CKD** |  |  |  |  |
| No | Regularly Active | 0.76 (0.63-0.93) | 0.008 | 0.312 |
| Yes | Regularly Active | 0.49 (0.17-1.39) | 0.179 | 0.312 |
| No | Weekend Warrior | 0.84 (0.72-0.98) | 0.026 | 0.312 |
| Yes | Weekend Warrior | 0.95 (0.53-1.68) | 0.847 | 0.312 |
| **T2DM** |  |  |  |  |
| No | Regularly Active | 0.76 (0.62-0.94) | 0.010 | 0.408 |
| Yes | Regularly Active | 0.63 (0.3-1.32) | 0.220 | 0.408 |
| No | Weekend Warrior | 0.85 (0.73-0.99) | 0.043 | 0.408 |
| Yes | Weekend Warrior | 0.78 (0.47-1.31) | 0.349 | 0.408 |
| **Hypertension** |  |  |  |  |
| Yes | Regularly Active | 0.73 (0.55-0.96) | 0.024 | 0.682 |
| No | Regularly Active | 0.76 (0.58-1.01) | 0.055 | 0.682 |
| Yes | Weekend Warrior | 0.94 (0.77-1.15) | 0.566 | 0.682 |
| No | Weekend Warrior | 0.74 (0.59-0.92) | 0.008 | 0.682 |

RCS analysis revealed a nonlinear inverse association between MVPA volume and gout risk in Model 1 and 2 (P-overall < 0.001; P-nonlinearity < 0.05; Fig. 5A-B). Notably, this protective association persisted in fully adjusted model (Model 3: P-overall = 0.010; Fig. 5C), though nonlinearity was attenuated (P-nonlinearity = 0.088). Further exclusion of individuals with MVPA >95th percentile yielded comparable dose-response curves (Fig. 5D-F).

Collectively, these findings demonstrate that total volume of MVPA—rather than its temporal distribution—is the primary determinant of reduced gout risk.

### 3. Inverse Probability of Treatment Weighting (IPTW) Analysis

To enhance the robustness of our findings, we further employed IPTW analysis. Covariate balance across key factors was effectively achieved, as evidenced by a substantial reduction in the absolute standardized mean differences after IPTW adjustment for both the RA vs. Inactive and WW vs. Inactive comparisons (Fig. 3A-B, Table S2-3). Following IPTW adjustment, the RA group showed a significantly lower gout risk compared to the Inactive group (Model 3: HR = 0.76, 95% CI: 0.62–0.93; P = 0.007; Fig. 3C). Similarly, the WW group also had a lower gout risk (Model 3: HR = 0.85, 95% CI: 0.72–0.99; P = 0.039; Fig. 3C). Weighted incidence rates were 0.91 vs. 1.19 per 1,000 person-years for RA vs. Inactive and 1.10 vs. 1.28 per 1,000 person-years for WW vs. Inactive (Table S4). After adjusting for total MVPA, no significant difference was found between WW and RA (Fig. 3D).

**Figure 3:**
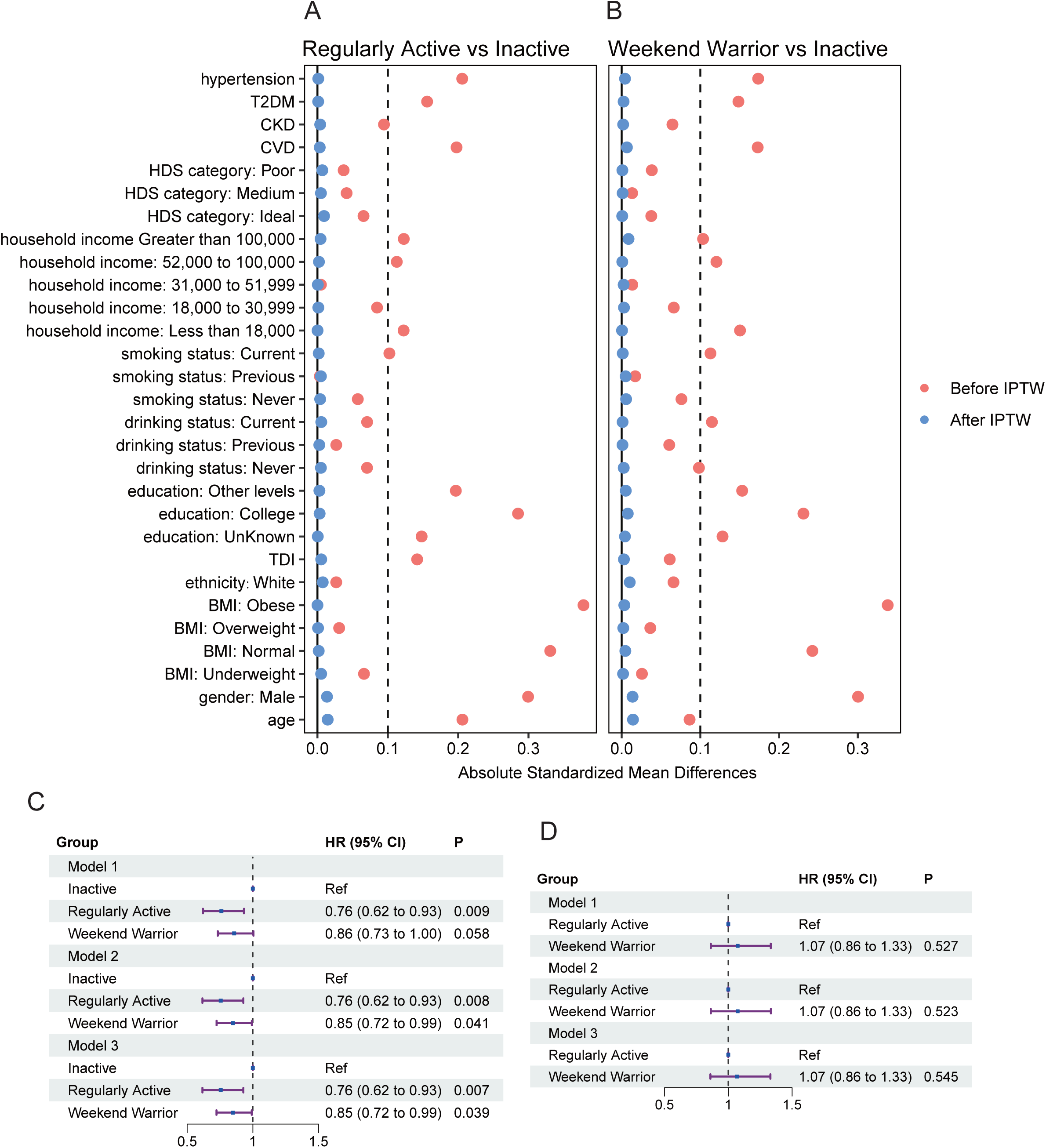
IPTW analysis of physical activity patterns and gout risk. (A) SMD for covariates between RA and Inactive groups before and after IPTW adjustment. (B) SMD for covariates between WW and Inactive groups before and after IPTW. (C) Forest plot of IPTW-adjusted HRs and 95% CIs for incident gout risk comparing RA and WW patterns to the Inactive group. (D) Forest plot of IPTW-adjusted HRs (95% CIs) comparing WW vs. RA patterns, after additional adjustment for total MVPA duration in all three models. The vertical dashed line represents HR = 1. IPTW: Inverse Probability of Treatment Weighting; SMD: absolute standardized mean differences

Overall, IPTW-weighted results support the primary findings: greater MVPA is associated with reduced gout risk, and both WW and RA patterns offer similar benefits when total MVPA volume is considered.

### 4. Role of PRS in the Association of Physical Activity and Gout Risk

Participants were stratified into tertiles representing low (bottom 1/3), middle (middle 1/3), and high genetic risk (top 1/3) based on their gout PRS. We then assessed the combined effects and interaction between physical activity patterns and PRS tertiles on gout risk (Fig. 4). Both RA and WW groups showed significantly reduced gout risk compared to inactive individuals in all three models. Importantly, the lack of significant interaction in all models (all P for interaction > 0.1) indicates that this protective effect of both physical activity patterns was consistently observed irrespective of whether individuals had low, middle, or high PRS. Collectively, these results indicate that higher PRS increases gout risk, while both RA and WW offer significant protection independent of genetic predisposition, with no evidence that genetic risk modifies the benefit of physical activity.

**Figure 4:**
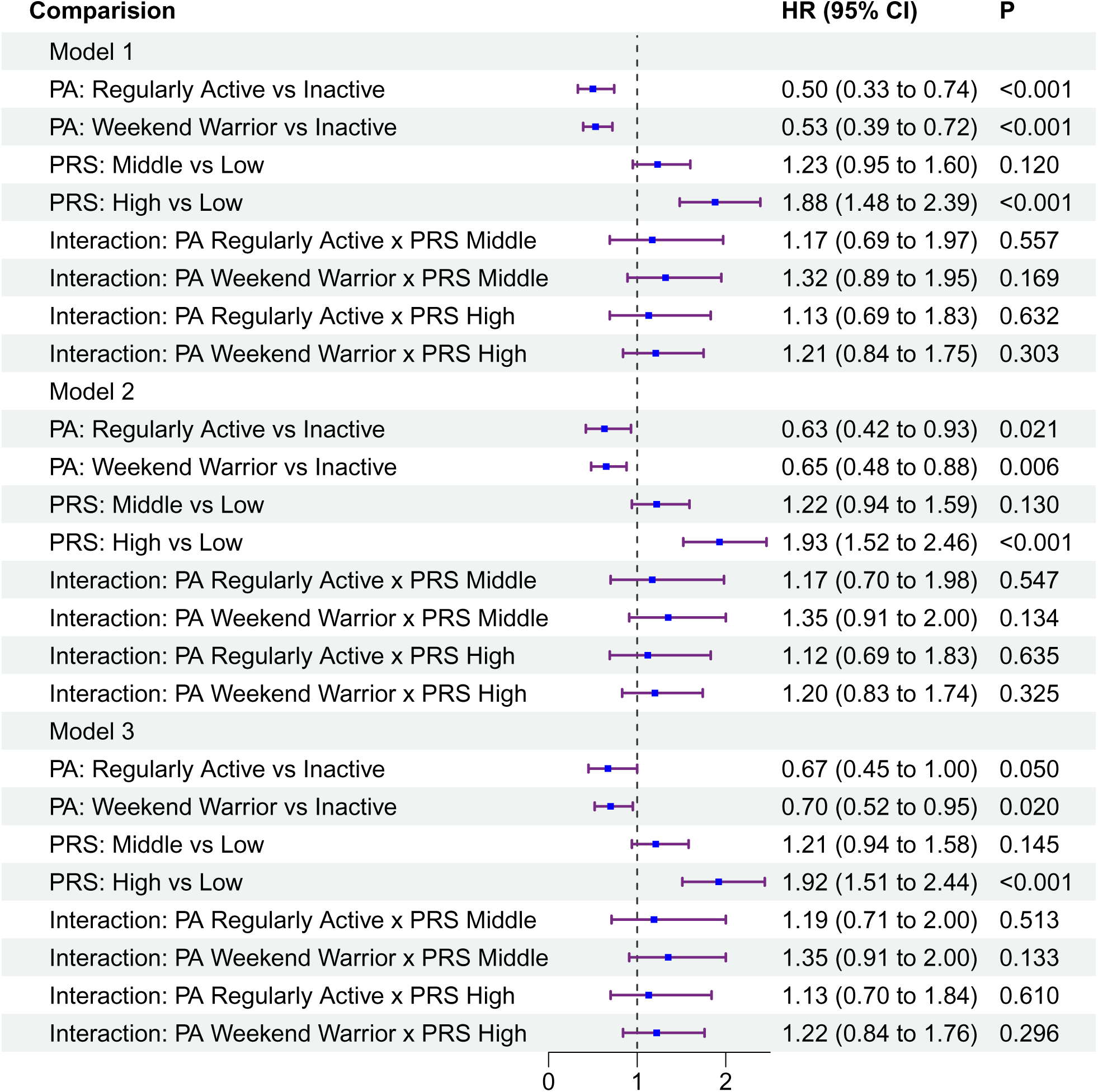
Associations of physical activity patterns with incident gout risk by genetic predisposition. Forest plot displaying HRs and 95% CIs for the association of physical activity patterns (RA, WW, Inactive [reference]) and gout PRS tertiles (Low [reference], Middle, High) with incident gout risk. The vertical dashed line represents the null effect (HR = 1). PRS, polygenic risk score.

**Figure 5:**
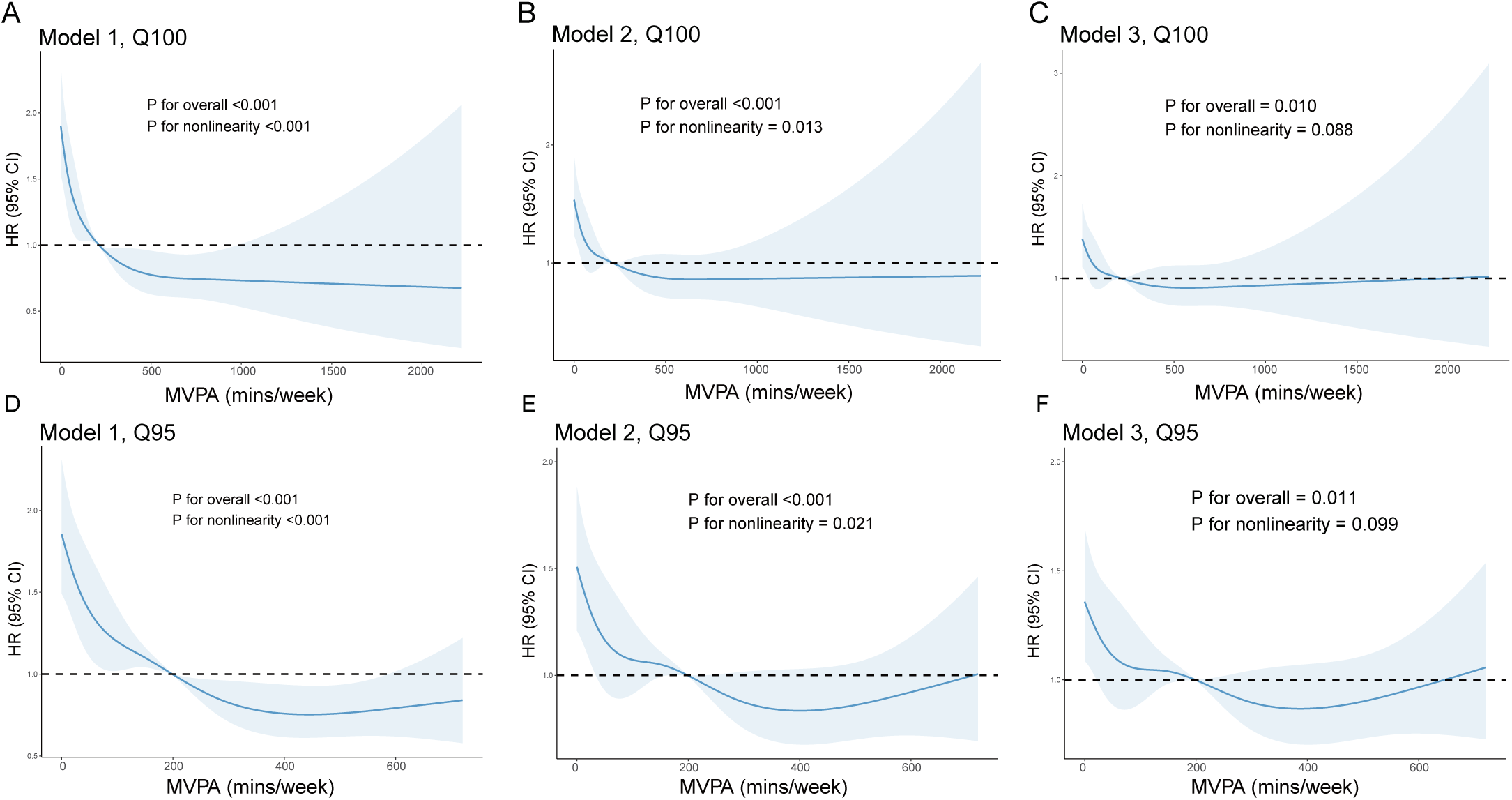
Relationship between MVPA volume and gout risk using RCS models. (A) RCS curve showing the multivariable-adjusted HRs (solid line) and 95% Confidence Intervals (shaded area) for incident gout across total weekly MVPA volume (reference = medium of total MVPA, dashed line at HR=1), adjusted for age and sex only (Model 1). (B) RCS curve adjusted as per Model 2 (additional covariates). (C) RCS curve adjusted as per Model 3 (full covariates). (D-F) Corresponding RCS curves (D: Model 1, E: Model 2, F: Model 3) after excluding individuals with MVPA volume > 95th percentile. P-overall and P-nonlinearity values are indicated for each model. RCS, Restricted Cubic Spline.

## Sensitivity Analyses

To assess the robustness of our findings, we conducted a series of sensitivity analyses. First, we varied the threshold used to define the “active” group based on MVPA duration, using the 25th (94.1 min/week), 50th (211.1 min/week), and 75th (374.4 min/week) percentiles of participant MVPA as alternative cut-offs. Second, we applied a more stringent definition of the WW pattern, requiring not only total MVPA ≥ 150 min/week but also that at least 75% of MVPA time was accumulated within 1-2 days. Third, to address potential reverse causation—specifically, the possibility that undiagnosed early gout might limit physical activity capacity) —we excluded participants who developed gout within 2 years of baseline (n = 249).

The results across these sensitivity analyses were consistent with our primary findings. Higher levels of MVPA remained significantly associated with a reduced risk of incident gout, while physical inactivity consistently emerged as a significant risk factor for gout development.

## Discussion

Based on a large prospective cohort from the UK Biobank, this study systematically investigated the association between different physical activity patterns and the risk of incident gout with leveraging objectively measured physical activity data from wrist-worn accelerometers. Our primary finding is that both the RA and WW pattern were independently associated with a significantly lower risk of developing gout compared to physical inactivity, even after extensive adjustment for potential confounding factors including BMI, gender, lifestyle, socioeconomic status, and comorbidities. After adjustment for total weekly MVPA minutes, no significant difference in gout risk reduction occurred between these two distinct temporal patterns of achieving the MVPA guidelines. This compellingly suggests that the total volume of MVPA achieved per week is the principal driver of the observed risk reduction for gout, not the specific timing or distribution of activity across the week.

Currently, high-quality research elucidating the impact of physical activity on gout risk remains limited [21,22]. This study utilized objectively measured physical activity data to demonstrate that RA and WW confer comparable protective effects against gout incidence. Our findings highlight the inherent protective role of physical activity in reducing gout risk and address the recall bias commonly associated with questionnaire-based studies [23,24]. Consequently, these results support the promotion of physical activity as a public health strategy for the primary prevention of gout, offering flexible options tailored to individuals with varying lifestyles.

Consistent with previous researches, our findings align with evidence showing similar protective effects of RA and WW patterns for cardiovascular disease, type 2 diabetes, and neurological disorders [25–27]. This consistency across health outcomes reinforces the concept that “dose,” measured as cumulative volume of activity above a moderate intensity, is paramount.

The persistence of the protective association after comprehensive adjustment, including for BMI, gender, comorbidities (CVD, CKD, T2DM, hypertension), and socioeconomic factors, strengthens the evidence for a potential causal relationship between MVPA and reduced gout risk. This was further bolstered by our IPTW analysis, which yielded consistent and statistically significant hazard ratios for both RA and WW patterns compared to Inactivity while effectively balancing baseline covariates. The robustness of the findings was confirmed across multiple sensitivity analyses, and extensive subgroup analyses showing no significant effect modification. Notably, no subgroup demonstrated heterogeneity in the benefit of being active (either pattern) versus inactive.

Our stratified analyses incorporating a validated gout PRS revealed a significant and clinically important layer of understanding. While genetic predisposition substantially increased the absolute risk of gout, as expected, the relative reduction in gout risk conferred by both RA and WW patterns was remarkably consistent across all genetic risk strata. There was no evidence of a statistically significant interaction between genetic susceptibility and physical activity pattern effectiveness. This indicates that engaging in adequate MVPA, irrespective of pattern, offers significant protection against gout even among individuals genetically predisposed to develop the condition. This finding underscores the broad applicability of physical activity as a preventive strategy for gout across different genetic backgrounds.

Our study findings have significant public health and clinical implications. They strongly support the inclusion of physical activity as a cornerstone for gout primary prevention strategies. Crucially, they suggest flexibility in implementation: while consistent, regular activity is beneficial, achieving the recommended MVPA volume primarily over weekends offers comparable protection against gout. This WW pattern might be a particularly feasible and sustainable option for individuals with time constraints or schedules that preclude weekday exercise. Such flexibility could significantly improve adherence to activity recommendations in the population. While weight loss remains paramount, this study provides robust evidence to warrant explicit recommendations regarding physical activity volume in future gout management and prevention guidelines, addressing a notable gap in the current 2020 ACR guidelines [7].

Limitations: First, accelerometer data were collected during a single 7-day period. While this provides an objective snapshot, it may not perfectly reflect an individual’s typical long-term activity pattern. Second, despite adjusting for numerous potential confounders and employing IPTW, residual confounding from unmeasured factors (e.g., undiagnosed renal impairment not captured in baseline history, or very specific dietary components like purine intake not captured by HDS) is always possible. Third, the study population consisted predominantly of individuals of White European ancestry, potentially limiting generalizability to other racial or ethnic groups where gout epidemiology and associated factors may differ.

## Conclusions

Engaging in moderate to vigorous physical activity (MVPA)—whether through regular routines or a “weekend warrior” pattern—significantly reduces the risk of developing gout compared to inactivity. This protective effect is primarily driven by the total volume of MVPA, not its temporal distribution pattern. Importantly, this benefit of adequate MVPA holds across diverse demographic subgroups and genetic risk strata. These findings highlight total physical activity volume as a key modifiable factor for gout prevention and support flexible approaches to achieving physical activity goals. Future gout prevention guidelines may explicitly incorporate recommendations regarding target MVPA volumes.

## Data Availability

Access requires registration and approval through the institutional access portal at www.ukbiobank.ac.uk/register-apply. Analytic code supporting this research will be available upon reasonable request to the corresponding author.

## Acknowledgements

This research was carried out using data from the UK Biobank (Application ID 545732). We are deeply grateful to all the participants who generously contributed their time and information to the UK Biobank. We also extend our appreciation to the dedicated teams involved in the design, implementation, and ongoing management of this invaluable resource.

## Code availability

Analytic code supporting this research will be available upon reasonable request to the corresponding author.

## Data availability

This work employs data from UK Biobank (Application ID 545732), a population-scale biomedical database. Access requires registration and approval through the institutional access portal at www.ukbiobank.ac.uk/register-apply.

